# Failing our Most Vulnerable: COVID-19 and Long-Term Care Facilities in Ontario

**DOI:** 10.1101/2020.04.14.20065557

**Authors:** David N. Fisman, Isaac Bogoch, Lauren Lapointe-Shaw, Janine McCready, Ashleigh R. Tuite, the COVID-19 Ontario Modelling Group

**Author notes:** Corresponding author: David Fisman, MD MPH FRCP(C), Room 686, 155 College Street, Toronto, Ontario, M5T 3M7. **Competing interests:** none.

## Abstract

**Background:** The COVID-19 epidemic has taken a fearsome toll on individuals residing in long-term care facilities (LTC). As of April 10, 2020 half of Canada’s COVID-19 deaths had occurred in LTC. We sought to better understand trends and risk factors for COVID-19 death in LTC in Ontario.

**Methods:** We analyzed a COVID-19 outbreak database created by the Ontario Ministry of Health, for the period March 29-April 7, 2020. Mortality incidence rate ratios for LTC were calculated with community living Ontarians aged > 69 used as the comparator group. Count-based regression methods were used to model temporal trends and identify associations between infection risk in staff and residents, and subsequent LTC resident death.

**Results:** Confirmed or suspected cases of COVID-19 were identified in 272/627 LTC by April 7, 2020. The incidence rate ratio for COVID-19 death was 13.1 (9.9-17.3) relative to community living adults over 69. Incidence rate ratio increased over time and was 87.28 (90% CrI 9.98 to 557.08) by April 7, 2020. Lagged infection in staff was a strong predictor of death in residents (e.g., adjusted IRR for death per infected staff member 1.17, 95% CI 1.11 to 1.26 at a 6-day lag).

**Interpretation:** Mortality risk in elders in Ontario is currently concentrated in LTC, and this risk has increased sharply over a short period of time. Early identification of risk requires a focus on testing and provision of personal protective equipment to staff, and restructuring the LTC workforce to prevent movement of COVID-19 between LTC.

**Funding:** The research was supported by a grant to DNF from the Canadian Institutes for Health Research (2019 COVID-19 rapid researching funding OV4-170360).

## Introduction

Communicable diseases do not respect boundaries. But they also do not affect all members of our society equally. Since its initial recognition in January 2020, it has been starkly apparent that SARS-CoV-2 disproportionately harms the most vulnerable in our population. Mortality due to COVID-19 is highest in the elderly and those with underlying medical conditions (1). In Canada, we had an early indication of the particular vulnerability of those living in long-term care (LTC). The first confirmed death in the country occurred on March 9^th^ 2020 in a resident of a North Vancouver care home (2).

Since then, outbreaks in LTC have been identified across the country. As of April 9 2020 nearly 50% (198/401) of all COVID-19 attributable deaths in the country have occurred in residents of LTC (3). In addition to the age and comorbidity profiles of residents, other factors make LTC especially susceptible to infectious disease outbreaks, including outbreaks of COVID-19 infection (4). These include: a lack of access to testing and personal protective equipment, the close quarters of residents, the difficulty of maintaining physical distancing among mobile patients with dementia, and a precariously employed workforce that can transmit the virus across LTC sites (5-7).

Recognizing and quantifying the scope of the issue and identifying which resident and worker factors may contribute to sudden increases in deaths is an important first step for ensuring that comprehensive policies and response measures are in place to protect residents and the health care workers who provide essential services to these residents. We sought to evaluate the risk of death among residents of LTC and identify risk factors associated with mortality, using data from the province of Ontario.

## Methods

Data for this study were obtained from the Ontario Ministry of Health and Long-term care as part of the province’s emergency “modeling table”. This included the following: (i) age and date of death of COVID-19-positive Ontario residents and (ii) cumulative death and COVID-19 positive case counts by date among LTC residents and staff. The available time series for LTC covered the 10-day period between March 29 and April 7, 2020, inclusive, however deaths prior to March 29 were included as part of the cumulative counts. The provincial death data included the time period from March 3 to April 11, 2020. Daily deaths and cases were estimated by taking the difference of cumulative cases and deaths on sequential days.

Population denominators for non-LTC deaths were derived from Statistics Canada estimates for the appropriate age group. Population denominators were not available for LTC facilities and were approximated as the total number of facility beds in Ontario (79,498) assuming complete occupancy. Age data for deaths reported in LTC were not available, but data show that approximately 93% of residents in LTC are aged 65 or older (8). In our main analysis, we estimated incidence rate ratios for COVID-19 deaths in the LTC resident population, compared to deaths in Ontario population aged over 70 years. In sensitivity analyses we compared outcomes in LTC residents to COVID-19 deaths in the entire Ontario population, the population aged >59 years, or >79 years. Cumulative deaths at the end of the respective time series were considered, with risk in both LTC residents and non-LTC residents considered to have begun on March 3, 2020. However, person-time denominators were adjusted for the fact that the LTC database ended 4 days before the provincial death database (i.e., 36 days in the LTC population and 40 days in the general Ontario population).

We also created a negative binomial regression model to evaluate the change in incidence rate ratio in LTC residents, as compared to the non-LTC population, during the period from March 29-April 7, 2020. Negative binomial regression was performed as a deviance statistic test suggested the data were not Poisson-distributed. Non-LTC residents aged > 69 were used as the comparator population, and model offsets were the population denominators described above. The model included three covariates: LTC residence, time (centered around April 3, 2020) and an interaction term of LTC residence (yes/no) multiplied by time. Model predictions were generated both forwards and backwards to generate graphical representations of how risk in the LTC population has changed over time.

Lastly, we created Poisson regression models that evaluated risk of death within LTC facilities as a function of the number of laboratory-confirmed, infected residents and confirmed infected staff at lags from 0-7 days, adjusting for date. As our model could not accommodate multiple simultaneous lags, we constructed 8 models, each of which included cumulative resident and staff infection as at identical lags (0 to 7 days. Deviance statistics indicated that counts were Poisson-distributed. LTC beds were used as model offsets. and confidence intervals were adjusted for clustering by LTC facility.

The study was approved by the Research Ethics Board of the University of Toronto.

## Results

A total of 627 LTC were included in the provincial dataset; of these 272 (43.4%) were identified as having either confirmed or suspected COVID-19 infection in residents or staff. No significant differences between LTC with and without confirmed COVID-19 infections were seen in number of licensed bed size, operator (e.g., for-profit vs. not-for-profit), or geographic location in Ontario (**Table 1**).

**Table 1.**
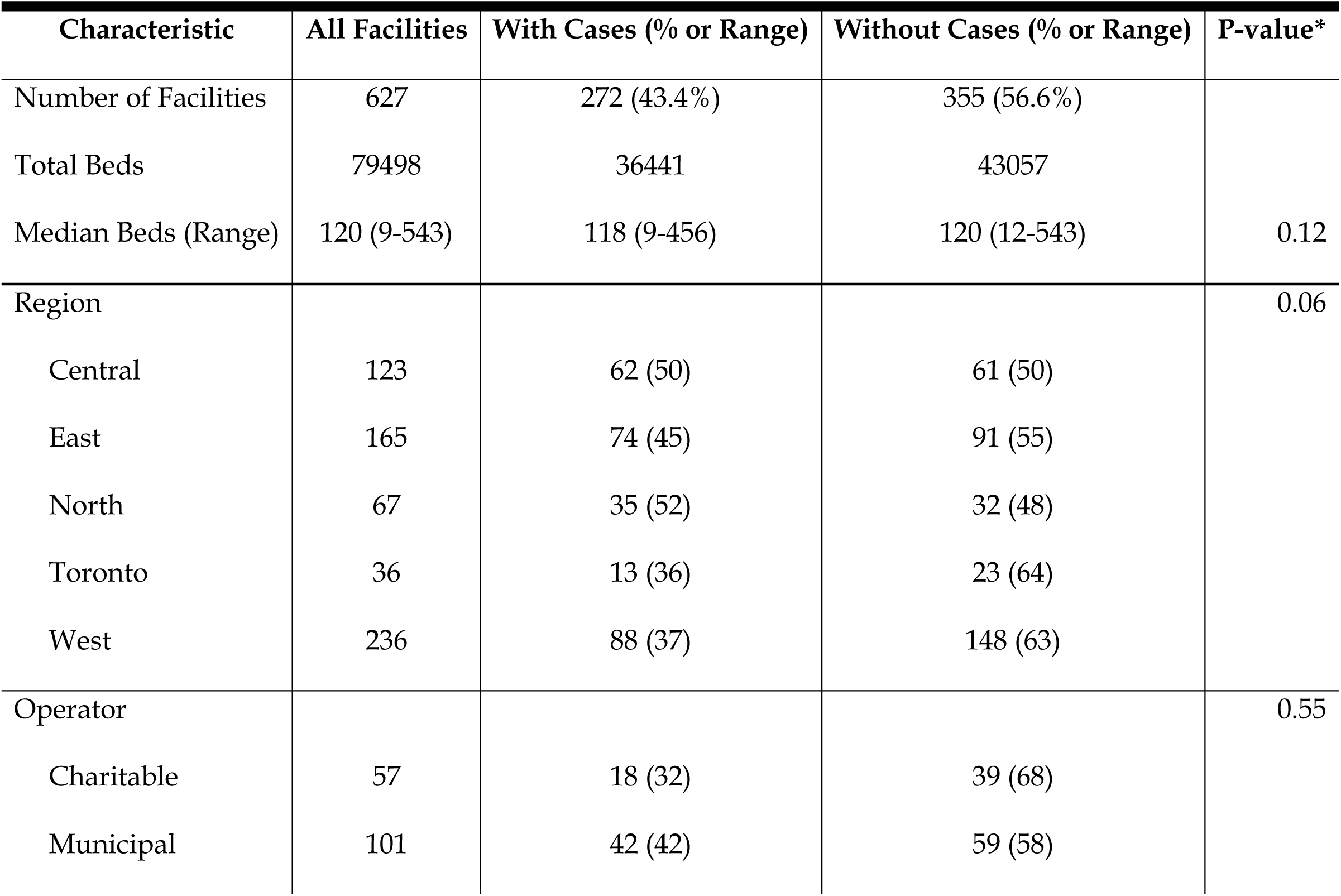
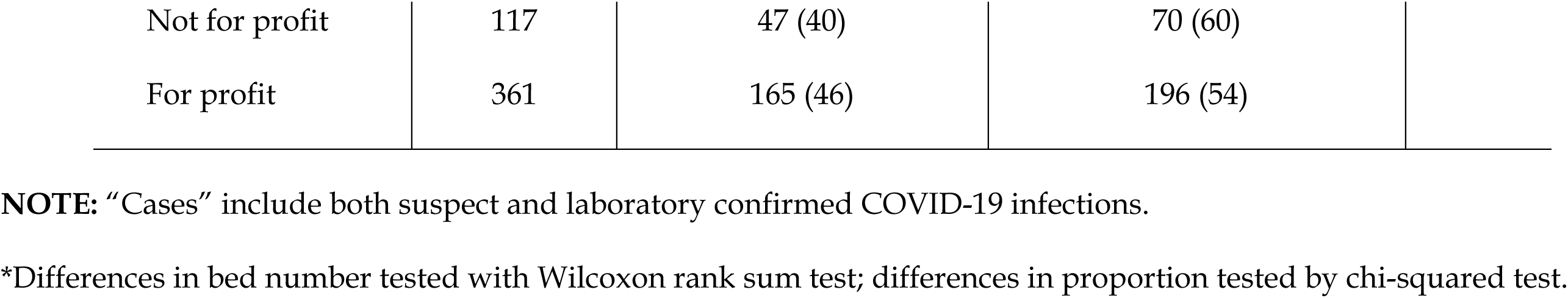
Characteristics of Ontario Long Term Care Facilities Included in Database.

The incidence of death due to COVID-19 was 13-fold higher in the LTC population than in Ontario residents aged > 69 years. When the whole population was used as the referent, the IRR for death was > 90 in this population; incidence was 23-fold higher when compared to those aged > 59 years, and 8-fold higher when compared to those those aged 80 and over not resident in LTC (**Table 2**).

**Table 2.** Incidence Rate Ratio for COVID-19 Mortality in LTC Residence. Using mortality data reported to April 11, 2020. A total of 83 deaths were reported in long-term care facilities to April 7, 2020 among 79,498 residents.

| **Comparator Population** | **Total Deaths** | **Comparator**<br>**Population Size** | **Incidence Rate Ratio**<br>**(95% CI)** |
| --- | --- | --- | --- |
| All ages | 269 | 14,566,547 | 90.4 (68.9 – 117.6) |
| 60 years and older | 252 | 3,447,723 | 23.1 (17.6-30.2) |
| 70 years and older | 229 | 1,731,315 | 13.1 (9.9 – 17.3) |
| 80 years and older | 169 | 642,571 | 7.6 (5.5 – 10.4) |

We identified significant interaction between time and risk associated with LTC residence (**Figure 1**). While risk of death in those not resident in LTC declined non-significantly over time, the rate ratio for death in LTC residents rose sharply, from 8.03 (90% CI 2.73 to 20.42) on March 29 to 87.28 (90% CI 9.98 to 557.08) by April 7, 2020.

In analyses focussed risk for death within LTC (**Table 2, Figure 2**) we found that lagged infections in institution staff were the strongest predictors of death in residents (**Figure 2**) and were significant at all lags (0 to 7 days) after adjustment for date and numbers of infected residents. The strongest effects were seen with infected staff at a 2 day lag (relative increase in death per infected staff member 20%, 95% CI 14-26%) and a 6 day lag (17%, 95% CI 11%-26%). By contrast the association between infection in residents and subsequent resident death was variable, and far weaker than the effect seen for staff, and was statistically significant only at a zero-day lag (increased risk per infected resident 8%, 95% CI 1% to 15%).

## Discussion

In this analysis we document the rapid spread of COVID-19 through Ontario’s LTC system, with a marked increase in risk of death among frail older residents in a brief period from late March to early April 2020. Issues such as crowding, use of communal space, low staffing ratios, and high care needs (with resultant high density of physical contact between residents and staff) have long been recognized as key drivers of vulnerability to outbreaks in the LTC setting (4, 7). In the context of COVID-19 this vulnerability has proven particularly deadly, with (as we demonstrate here), an incidence of mortality more than 13 times higher than that seen in community living elders over age 69 during a similar period, with relative risk of death rising sharply over time.

We find also that documented infection in facility staff, as opposed to residents, is a strong identifiable risk factor for mortality in residents, with temporality suggesting that residents are infected by staff, and not vice versa. Although it might be argued that the limited nature of testing, and a tendency to test staff and residents for COVID-19 after a resident dies might lead to spurious associations between identified infections and deaths, the temporality of association and differential strength of effect in staff and residents suggests that this effect may be causal. The greater mobility and connectedness of staff in the LTC setting, as opposed to residents, lends biological plausibility to this effect (9).

Transmission of infection is not the only mechanism whereby infection in staff could result in increased mortality in a vulnerable older population. Fear of COVID-19 may result in absenteeism by staff, which could itself lead to death through dehydration and other mechanisms in a high-needs population, which would be consistent with the lagged effects we demonstrate here. Such tragedies have been documented in Canada recently (10).

The prevention of such deaths requires strategic guidance from health regions and the provision of sufficient testing and personal protective equipment. Provision of personal protective equipment has benefits both in bidirectional prevention of SARS-CoV-2 transmission, and in providing workers peace of mind, in order to stay on the job. Expanded testing, including testing of minimally symptomatic infection, will facilitate early identification of infection, and implementation of effective infection control strategies. Also needed are integrated regional approaches to LTC human resource management, such as limiting workers to a single facility and ensuring that these workers earn a living wage to prevent the need for multiple jobs while at the same time maintaining adequate staffing levels (11).

Like any observational study, this study has limitations, including possible incompleteness of data collected rapidly during an outbreak, inconsistency in testing across Ontario, and absence of individual-level data on LTC infections and deaths. We regard our outcome of interest, death in LTC from COVID-19 to be less likely misclassified than non-fatal infection in staff and residents. If misclassification of infection status in these individuals occurs at random, that would likely mean the effects reported here are lower bound effects. If under-identification of both fatal infections and infections in residents and staff are clustered by home, that would result in effects that are biased upwards. The temporality in the effects we observe provides a degree of reassurance in this regard.

## Conclusions

In summary, we document that the rapid movement of COVID-19 through Ontario’s LTC system has resulted in a marked surge in mortality in that population, as compared to community-living elders. We find evidence that links mortality to infection in LTC staff, highlighting the urgent need for improved infection control, more widespread testing, access to personal protective equipment, and economic protections and support for those who do this important work.

## Data Availability

Data are the property of the Government of Ontario and are not publicly available.

## Figure Legends

**Figure 1.** Model-based Estimation of COVID-19 Death Risk in Ontario. The two curves represent modeled deaths per 1000 individuals in the long-term care population (teal curve) and the community-dwelling population aged > 69 in Ontario (pink curve). Shaded areas represent 95% confidence intervals.

**Figure 2.** Incidence Rate Ratios for Death in Long Term Care, by Lagged Infections in Residents and Staff. Incidence rate ratios (IRR) from Poisson regression models evaluating lagged associations between confirmed COVID-19 staff infections (pink) and resident infections (green). Circles represent IRR, and vertical lines 95% confidence intervals. Elevated risk is seen with staff-person infections at all lags. Elevated risk is seen only with resident infections at 0-day lag (i.e., simultaneous with deaths).

